# Performance and Implementation Evaluation of the Abbott BinaxNOW Rapid Antigen Test in a High-throughput Drive-through Community Testing Site in Massachusetts

**DOI:** 10.1101/2021.01.09.21249499

**Authors:** Nira R. Pollock, Jesica R. Jacobs, Kristine Tran, Amber Cranston, Sita Smith, Claire O’ Kane, Tyler Roady, Anne Moran, Alison Scarry, Melissa Carroll, Leila Volinsky, Gloria Perez, Pinal Patel, Stacey Gabriel, Niall J. Lennon, Lawrence C. Madoff, Catherine Brown, Sandra C. Smole

## Abstract

**Background:** Rapid diagnostic tests (RDTs) for SARS-CoV-2 antigens (Ag) that can be performed at point-of-care (POC) can supplement molecular testing and help mitigate the COVID-19 pandemic. Deployment of an Ag RDT requires an understanding of its operational and performance characteristics under real-world conditions and in relevant subpopulations. We evaluated the Abbott BinaxNOW™ COVID-19 Ag Card in a high-throughput, drive-through, free community testing site in Massachusetts (MA) using anterior nasal (AN) swab RT-PCR for clinical testing.

**Methods:** Individuals presenting for molecular testing in two of seven lanes were offered the opportunity to also receive BinaxNOW testing. Dual AN swabs were collected from symptomatic and asymptomatic children (≤ 18 years) and adults. BinaxNOW testing was performed in a testing pod with temperature/humidity monitoring. One individual performed testing and official result reporting for each test, but most tests had a second independent reading to assess inter-operator agreement. Positive BinaxNOW results were scored as faint, medium, or strong. Positive BinaxNOW results were reported to patients by phone and they were instructed to isolate pending RT-PCR results. The paired RT-PCR result was the reference for sensitivity and specificity calculations.

**Results:** Of 2482 participants, 1380 adults and 928 children had paired RT-PCR/BinaxNOW results and complete symptom data. 974/1380 (71%) adults and 829/928 (89%) children were asymptomatic. BinaxNOW had 96.5% (95% confidence interval [CI] 90.0-99.3) sensitivity and 100% (98.6-100.0) specificity in adults within 7 days of symptoms, and 84.6% (65.1-95.6) sensitivity and 100% (94.5-100.0) specificity in children within 7 days of symptoms. Sensitivity and specificity in asymptomatic adults were 70.2% (56.6-81.6) and 99.6% (98.9-99.9), respectively, and in asymptomatic children were 65.4% (55.6-74.4) and 99.0% (98.0-99.6), respectively. By cycle threshold (Ct) value cutoff, sensitivity in all subgroups combined (n=292 RT-PCR-positive individuals) was 99.3% with Ct ≤25, 95.8% with ≤30, and 81.2% with ≤35. Twelve false positive BinaxNOW results (out of 2308 tests) were observed; in all twelve, the test bands were faint but otherwise normal, and were noted by both readers. One invalid BinaxNOW result was identified. Inter-operator agreement (positive versus negative BinaxNOW result) was 100% (n = 2230/2230 double reads). Each operator was able to process 20 RDTs per hour. In a separate set of 30 specimens (from individuals with symptoms ≤7 days) run at temperatures below the manufacturer’s recommended range (46-58.5°F), sensitivity was 66.7% and specificity 95.2%.

**Conclusions:** BinaxNOW had very high specificity in both adults and children and very high sensitivity in newly symptomatic adults. Overall, 95.8% sensitivity was observed with Ct ≤ 30. These data support public health recommendations for use of the BinaxNOW test in adults with symptoms for ≤7 days without RT-PCR confirmation. Excellent inter-operator agreement indicates that an individual can perform and read the BinaxNOW test alone. A skilled laboratorian can perform and read 20 tests per hour. Careful attention to temperature is critical.

## Introduction

Nucleic acid amplification tests (NAAT) for SARS-CoV-2, the etiologic agent of COVID-19, can be highly sensitive and are being performed at high volumes in centralized laboratories around the world [1, 2]. Unfortunately, shortages of testing reagents and logistic barriers (e.g. sample transport) have impacted access to molecular testing and turnaround time for results. The need for decentralized testing options with short turnaround times has led to the development of rapid diagnostic tests (RDTs) for point-of-care (POC) use that detect SARS-CoV-2 nucleocapsid antigen (Ag) in as little as 15 minutes. As of December 26, 2020, there are 11 Ag RDTs with FDA Emergency Use Authorization (EUA) [3]. Most of these tests can be performed by personnel without formal laboratory training in patient care settings that operate minimally under a Clinical Laboratory Improvement Amendments (CLIA) Certificate of Waiver (COW).

Reported clinical sensitivities for Ag RDTs performed at POC in individuals suspected of COVID-19 vary widely, ranging from 74% to 97% compared with RT-PCR [4-12]. Reports of false negative and false positive Ag RDT results [13] have raised concerns regarding their use, although a range of settings would benefit from RDT availability (e.g. K-12 schools, nursing homes, community testing centers, and home settings). Gaps in knowledge of how Ag RDTs perform in asymptomatic individuals and children remain.

In symptomatic adults, viral loads in nasopharyngeal (NP) samples peak within the first week of symptom onset, then decrease over a variable time frame; RNA levels in asymptomatically infected adults appear to follow similar kinetics [14, 15]. Ag concentrations have been shown in one study to correlate tightly with Ct values in NP samples from adults and children [16]. While NP sampling remains the reference method, anterior nasal (AN) sampling substantially increases testing access and acceptability, and a recent head-to-head study suggested that Ag RDTs performed on paired NP/AN samples yielded similar results [5].

The BinaxNOW COVID-19 Ag CARD [8] has FDA EUA for AN swab samples and can provide visually-read results at POC in 15 minutes. The potential to use this test at large scale, combined with the lack of data for test performance in asymptomatic adults and in children, motivated us to perform an implementation and performance evaluation in a high-volume, high-prevalence community testing site currently using AN swab RT-PCR for clinical testing.

## Methods

### Study population

The study was performed between October 26-December 22, 2020 (all ages October 26-November 12, 2020, and children only from December 11-22, 2020), at the Lawrence General Hospital “Stop the Spread” drive-through testing site, which accommodates Massachusetts residents from the surrounding area. No study-specific effort was made to recruit individuals to present to the testing site. Two of seven drive-through lanes were utilized for the study. Verbal consent for dual AN swabbing was obtained from adults and guardians of minors (with verbal assent for ages 7-17). Participants were informed that they would be called for positive Ag RDT results only and that positive results would require isolation while waiting for the RT-PCR result. Presence or absence of symptoms (sore throat, cough, chills, body aches, shortness of breath, fever, runny nose, congestion, nausea, vomiting, diarrhea, loss of taste or smell) was recorded for each individual, including the date of symptom onset. Participants whose symptoms started on the day of testing were classified as Day 0. The study was reviewed by the Massachusetts Department of Public Health IRB and deemed not human subject research.

### Swab collection procedure

Cars with consented patients were marked with a glass marker, notifying the specimen collector to collect two AN swabs rather than one. Swab collection details are in Supplementary Methods; in brief, collection involved swabbing both nostrils with each swab and operators alternated which swab was collected first (for RT-PCR vs BinaxNOW). BinaxNOW swabs were captured in an empty sterile tube and taken to the testing pod by a designated “runner.” Time of sample collection was recorded.

### Abbott BinaxNOW performance

Details of kit storage, quality control, and testing and results reporting procedures are in Supplementary Methods.

### RT-PCR assay

Dry AN swabs were collected per site routine and transported at room temperature to the Broad Institute for testing using the CRSP SARS-CoV-2 Real-time Reverse Transcriptase (RT)-PCR Diagnostic Assay under EUA [17]. Details are in Supplementary Methods.

### Results reporting

All positive BinaxNOW results were reported to individuals the same day by Massachusetts Department of Public Health (DPH) epidemiologists. Participants with positive BinaxNOW results were informed that the result was presumptive and that they should maintain isolation for a minimum of 10 days based upon the result, but that a confirmatory result would be obtained by RT-PCR and reported in 1-2 days. If the RT-PCR result was negative, they could discontinue isolation. RT-PCR results were provided to the patient by the Lawrence General Hospital’s portal or by walk up to a designated location at the Lawrence General Hospital. RT-PCR results were reported to DPH through routine electronic laboratory reporting mechanisms and individuals with positive results were referred to local boards of health or the Community Tracing Collaborative for instruction on isolation and contact tracing.

### Statistical analysis

Sensitivity, specificity, negative predictive value (NPV), and positive predictive value (PPV) for the BinaxNOW were calculated using the RT-PCR result as the reference. 95% CI were calculated using the Clopper-Pearson method. Analyses utilized Microsoft Excel and GraphPad Prism.

## Results

Four different lots of BinaxNOW kits were used for the study. Each operator was able to set up a new test every 2-3 minutes, and two operators were able to manage testing of samples coming from two drive-through lanes. All tests were initiated within 1 hour of collection, per manufacturer instructions. Temperature and humidity in the testing pod (Supplementary Methods) between 7:30AM-6:00PM ranged from 33.3-75.7°F and 27.8-81.0%, respectively, in the shipping container and from 67.6-81.6°F and 20.8-69.0%, respectively, in the trailer. Tests for the main study were not run until the temperature reached 59°F, per manufacturer’s recommendation. Data for tests performed out of the temperature range in the package insert (<59°F) were analyzed separately (below).

Of 2482 participants [excluding samples tested at <59°F (n=94); inconclusive RT-PCR results (n=26); and missing data (n=54)], 2308 had paired PCR/BinaxNOW results and complete symptom data, including 829 asymptomatic children, 974 asymptomatic adults, 99 symptomatic children, and 406 symptomatic adults. Symptomatic individuals were further classified as ≤7 days versus >7 days of symptoms. Clinical data for the study population are presented in Table 1 (demographics) and Supplementary Tables 1 and 2 (symptoms).

**Table 1.** Clinical characteristics of adult and pediatric patients contributing paired samples.

|  | **Adults,**<br>**symptomatic**<br>**(n=407)** | **Adults,**<br>**asymptomatic**<br>**(n=974)** | **Children,**<br>**symptomatic**<br>**(n=99)** | **Children,**<br>**asymptomatic**<br>**(n=829)** |
| --- | --- | --- | --- | --- |
| **Age in years, n (%)** |  |  |  |  |
| <7 | n/a | n/a | 17 | 244 |
| 7-13 | n/a | n/a | 42 | 339 |
| 14-18 | n/a | n/a | 40 | 246 |
| 19-29 | 120 | 212 | n/a | n/a |
| 30-49 | 189 | 392 | n/a | n/a |
| 50-69 | 89 | 312 | n/a | n/a |
| 70 and older | 8 | 58 | n/a | n/a |
| **Sex, % female** | 58.9 | 56.4 | 61.6 | 52.0 |
| **Days of symptoms prior to**<br>**COVID test, median**<br>**(IQR)** | 3 (2,5)<sup>a</sup> | n/a | 2 (1,4)<sup>b</sup> | n/a |

<sup>a</sup>Range 0-55 days<sup>b</sup>Range 0-60 days

**Table 2.**
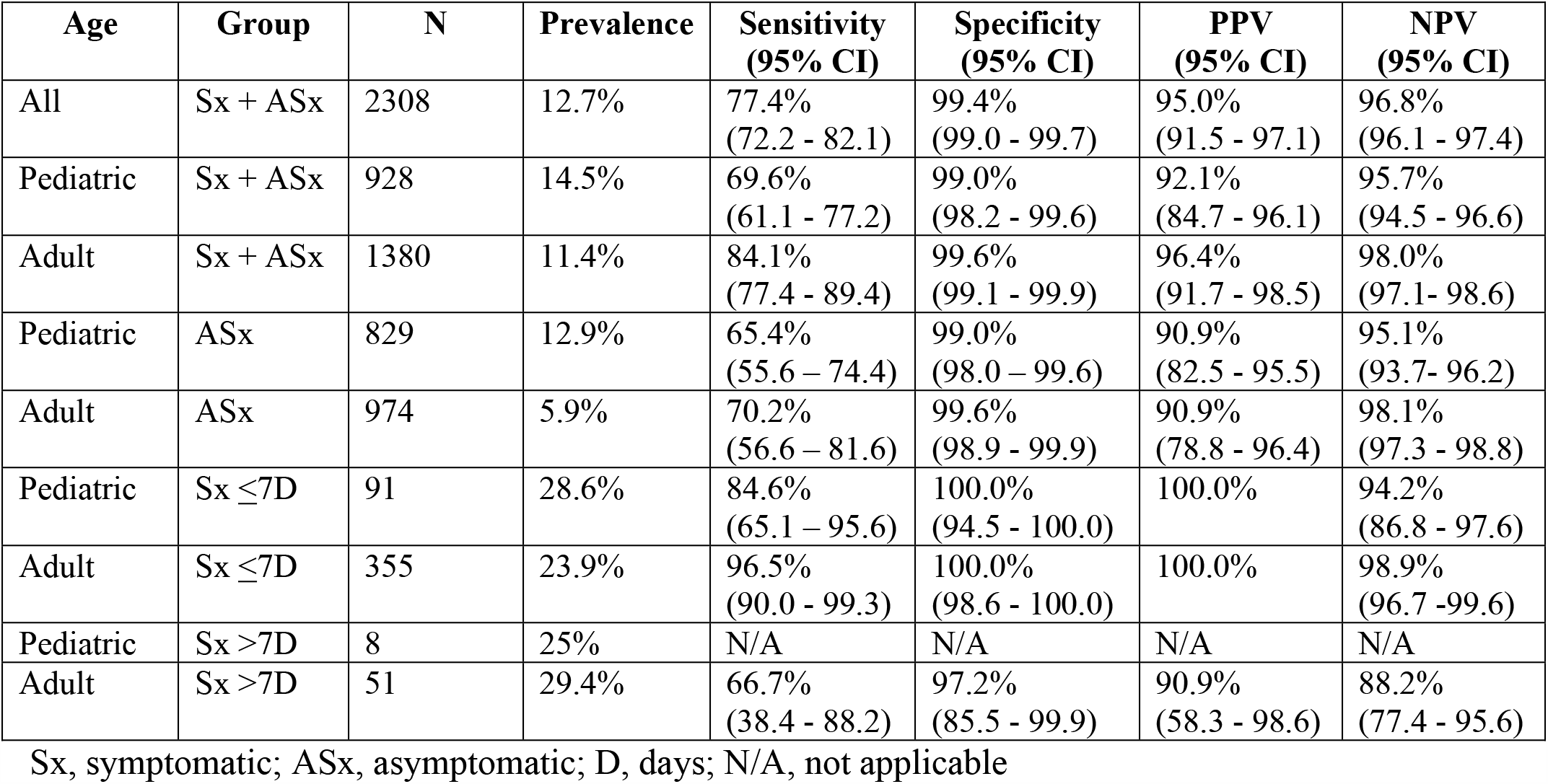
**Performance of the Abbott BinaxNOW versus RT-PCR (reference method) for detection of SARS-CoV-2 in anterior nasal swab samples from adult and pediatric (<18) patients**

| Age | Group | N | Prevalence | Sensitivity<br>(95% CI) | Specificity<br>(95% CI) | PPV<br>(95% CI) | NPV<br>(95% CI) |
| --- | --- | --- | --- | --- | --- | --- | --- |
| All | Sx + ASx | 2308 | 12.7% | 77.4%<br>(72.2 - 82.1) | 99.4%<br>(99.0 - 99.7) | 95.0%<br>(91.5 - 97.1) | 96.8%<br>(96.1 - 97.4) |
| Pediatric | Sx + ASx | 928 | 14.5% | 69.6%<br>(61.1 - 77.2) | 99.0%<br>(98.2 - 99.6) | 92.1%<br>(84.7 - 96.1) | 95.7%<br>(94.5 - 96.6) |
| Adult | Sx + ASx | 1380 | 11.4% | 84.1%<br>(77.4 - 89.4) | 99.6%<br>(99.1 - 99.9) | 96.4%<br>(91.7 - 98.5) | 98.0%<br>(97.1 - 98.6) |
| Pediatric | ASx | 829 | 12.9% | 65.4%<br>(55.6 - 74.4) | 99.0%<br>(98.0 - 99.6) | 90.9%<br>(82.5 - 95.5) | 95.1%<br>(93.7 - 96.2) |
| Adult | ASx | 974 | 5.9% | 70.2%<br>(56.6 - 81.6) | 99.6%<br>(98.9 - 99.9) | 90.9%<br>(78.8 - 96.4) | 98.1%<br>(97.3 - 98.8) |
| Pediatric | Sx $\leq 7$D | 91 | 28.6% | 84.6%<br>(65.1 - 95.6) | 100.0%<br>(94.5 - 100.0) | 100.0% | 94.2%<br>(86.8 - 97.6) |
| Adult | Sx $\leq 7$D | 355 | 23.9% | 96.5%<br>(90.0 - 99.3) | 100.0%<br>(98.6 - 100.0) | 100.0% | 98.9%<br>(96.7 - 99.6) |
| Pediatric | Sx $> 7$D | 8 | 25% | N/A | N/A | N/A | N/A |
| Adult | Sx $> 7$D | 51 | 29.4% | 66.7%<br>(38.4 - 88.2) | 97.2%<br>(85.5 - 99.9) | 90.9%<br>(58.3 - 98.6) | 88.2%<br>(77.4 - 95.6) |

Sx, symptomatic; ASx, asymptomatic; D, days; N/A, not applicable

### BinaxNOW performance in adults and children (≤18)

Sensitivity, specificity, positive predictive value (PPV) and negative predictive value (NPV) calculations for BinaxNOW results versus RT-PCR results as the reference, for each clinical subgroup, are presented in Table 2. Tables with data for each subgroup are presented in Supplementary Table 3.

Sensitivity in adults with symptoms ≤ 7 days was 96.5%, similar to that in the BinaxNOW package insert (97.1%) [8]. Sensitivity in children with symptoms ≤ 7 days was 84.6%. Specificity in both groups was 100%. Relative to symptomatic individuals, sensitivity in asymptomatic adults and children was lower at 70.2% and 65.4%, respectively, while specificity remained high (99.6%/99.0%, respectively). The anticipated NPV/PPVs with varying prevalence, based on the observed sensitivity/specificity in each subgroup, are presented in Supplementary Table 4.

### Discordant analysis and analysis of Ct values

There were 12 false positive BinaxNOW results across all 2308 individuals tested; 4 were in asymptomatic adults, 7 in asymptomatic children, and 1 in a symptomatic adult with >7 days of symptoms. All twelve positives were scored as faint bands by both independent readers, and there was nothing unusual noted about band morphology. 61/66 false negatives had dual read; 61/61 were scored as negative by both independent readers. There was no apparent correlation between false positives/false negatives and lot or kit number in testing through Nov. 12^th^. In the pediatric-only phase initiated Dec. 11^th^, all 6 false positive samples observed utilized one lot that was not used in the first phase; there was no correlation between false negatives and lot number.

Distributions of Ct values for RT-PCR positive symptomatic (by days post symptom onset) and asymptomatic children and adults are shown in Figure 1; false negative versus true positive paired BinaxNOW results are indicated for all individuals. As expected, false negative BinaxNOW results were paired with RT-PCR tests with higher Ct values. Ct distributions in the four main clinical subgroups are shown in Figure 2. Median Ct values (IQR) for children and adults symptomatic for ≤ 7 days were 24.2 (16.3-27.3) and 20.5 (17.2-26.5), respectively, and for asymptomatic children and adults were 26.8 (20.5-32.8) and 26.9 (20.6-32.8), respectively. Sensitivity was evaluated at three different Ct cutoffs: ≤25, ≤30, ≤35 (Supplementary Table 5). Sensitivity in all subgroups combined (n=292 RT-PCR-positive individuals) was 99.3% with Ct ≤25, 95.8% with ≤30, and 81.2% with ≤35. Band strength (1= faint, n = 41; 2 = medium, n = 15; 3 = strong, n = 170) as interpreted by the primary reader for the 226 true positive BinaxNOW tests correlated clearly with Ct value, with median Cts of 29.7 (27.6-32.7), 27.7 (25.5-29.1), and 19.5 (16.4-22.8), respectively, as shown in Supplementary Figure 1. All but one (65/66) false negative BinaxNOW results had a paired RT-PCR cycle threshold value >25, and 9 had Ct ≤30; the median (IQR) cycle threshold value for these false negatives was 33.7 (32.1-34.8). Distribution of the 66 false negative results among clinical subgroups is shown in the 2×2 tables in Supplementary Table 3.

**Figure 1.**
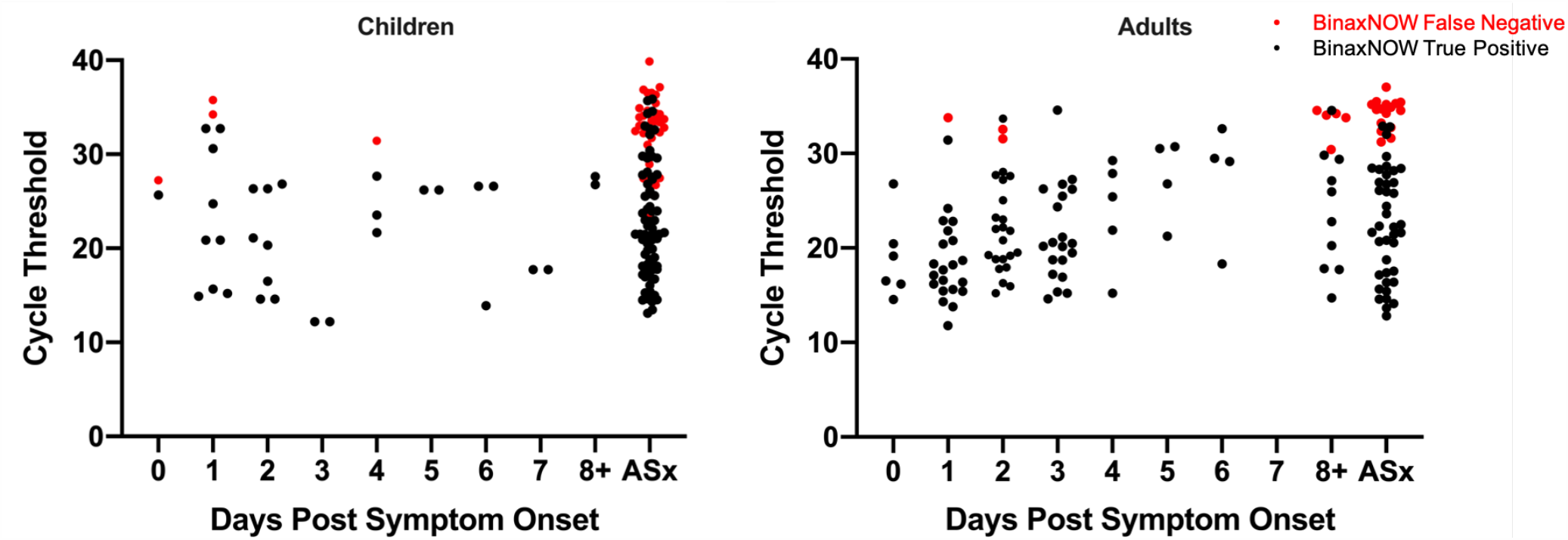
Distribution of Cycle Threshold (Ct) Values in RT-PCR-positive Children and Adults by Days Post Symptom Onset. Ct values for each RT-PCR-positive individual are shown; red circles indicate false negative BinaxNOW results and black circles, true positive BinaxNOW results. Participants whose symptoms started on the day of testing are indicated as Day 0. ASx, asymptomatic.

**Figure 2.**
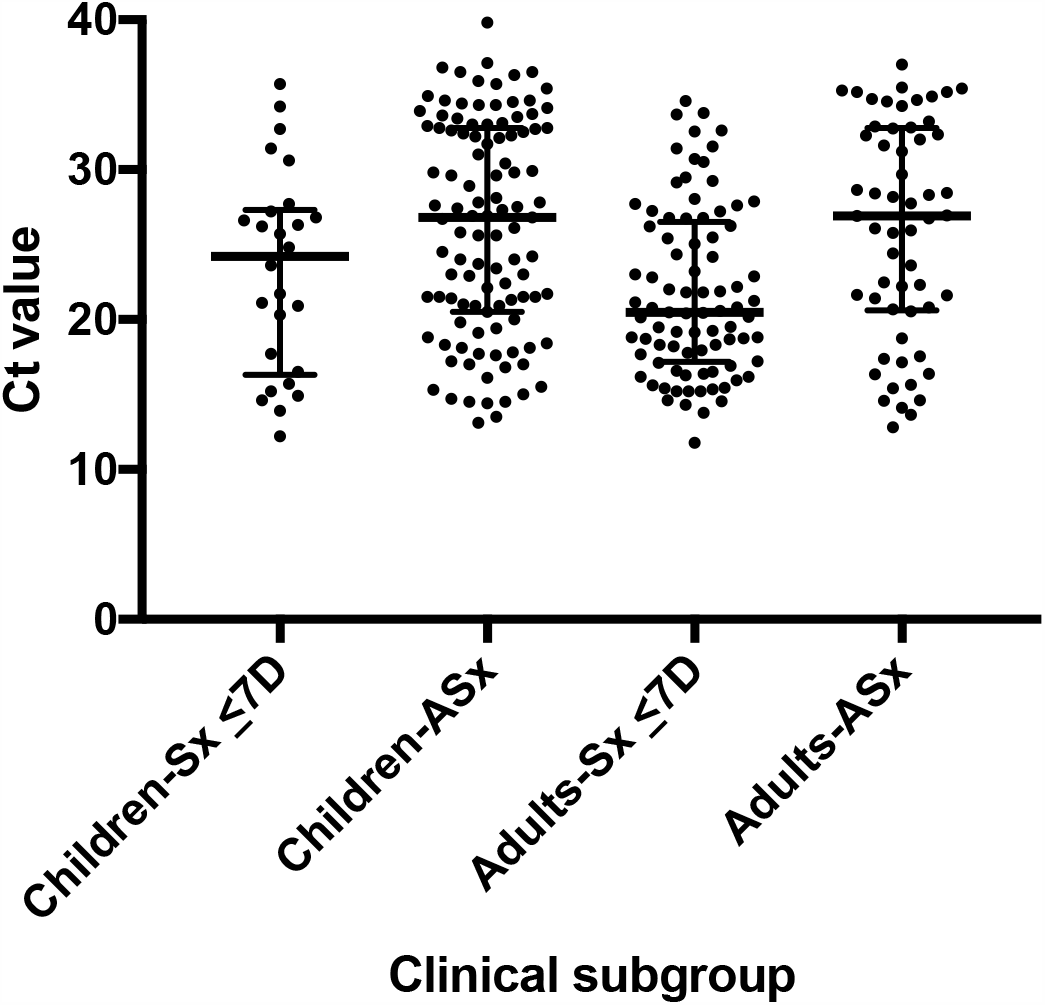
Distribution of Ct Values in RT-PCR-positive Individuals. Cycle threshold (Ct) values for RT-PCR positive individuals in each of four clinical subgroups are shown; horizontal bars represent median and IQR. Sx, symptomatic; ASx, asymptomatic; D, Days. Median Ct values (IQR) in Sx ≤ 7D: Children, 24.2 (16.3-27.3) and Adults, 20.5 (17.2-26.5) and in ASx: Children, 26.8 (20.5-32.8) and Adults, 26.9 (20.6-32.8).

### Operational findings

A small number of BinaxNOW tests were performed at temperatures below the manufacturer’s recommended temperature range due to difficulty maintaining temperature in the antigen testing location and interest in exploring the effect of low temperature on test performance. These data were analyzed separately from the main study. In 30 specimens from individuals (3 children and 27 adults) with symptoms ≤7 days run at low temperature (46-58.5°F), sensitivity was 66.7% and specificity 95.2% (Supplementary Table 6), in comparison to sensitivity of 93.7% and specificity of 100% in individuals with ≤7 days symptoms when testing was performed at >59°F. Relative humidity ranged from 45.7-66.4% during the two testing days with low temperatures (≤ 59°F). For false positive or false negative tests performed at >59 F, the humidity range was 27.2-62.6% and 22.6-61.5%, respectively, mirroring the overall humidity range and distribution.

Inter-operator agreement was excellent, with the two readers agreeing on the positive versus negative result for all 2230 BinaxNOW tests that had dual reads. The two readers disagreed on the strength of the positive band (faint vs medium vs strong) in 5 cases. Overall, readers noted that detection of faint positive bands required very close observation. While we did observe occasional inconsistent band intensity across the width of some test bands, we did not identify a correlation between that finding and false positivity. One invalid BinaxNOW result was identified during the study (a manufacturing issue whereby plastic covered the test strip, preventing the buffer from making contact with the test strip).

## Discussion

The development of Ag RDTs has expanded the options for POC COVID-19 testing and raised critical questions about how these tests could and should be used. Prominent examples of both false negative and false positive results [13] on Ag RDTs have raised questions about how well the tests perform in real world settings, even if performed carefully by trained operators. Major gaps in performance data, including performance in asymptomatic adults and both symptomatic and asymptomatic children, have generated uncertainty about how to optimally deploy Ag RDTs, and emerging comparison studies reiterate that not all Ag RDTs perform equivalently [4, 18]. The FDA EUA for the BinaxNOW RDT was based upon 102 subjects, all adults, who had symptomatic COVID-19 with symptom onset within 7 days prior to testing [8]. In order to understand how well the BinaxNOW RDT could perform in both symptomatic and asymptomatic adults and children, we implemented the test at a high-volume community testing site already experienced in collecting AN samples for RT-PCR, performing the test with careful attention to sample collection, test performance, results documentation, and quality control. The BinaxNOW was performed by trained laboratory personnel. This study design allowed us to evaluate performance of the BinaxNOW in a real-world but also best-case scenario, and to provide the first data on performance in a large population of children.

We found that test specificity was very high in this study in all populations, while sensitivity was variable. Sensitivity was very high in symptomatic adults within the first 7 days of illness, and less so in symptomatic children. Sensitivity in asymptomatic adults and children was even lower, corresponding with the broad viral load distribution observed in this population (likely capturing early and late infections given unknown disease onset). For all groups, sensitivity was highest in individuals with highest viral loads.

Our study yielded some important operational findings relevant to test implementation. Inter-operator agreement on positive/negative results was 100%, confirming that only one person is needed to read each test result. While one skilled operator was able to run 20 tests per hour, two people were needed to run tests in high volume, and additional people were needed to transport samples from the collection site to the testing site. We noticed a substantial impact of low temperature (below the manufacturer’s recommended range) on test accuracy, reinforcing that temperature is a critical parameter for test performance; the instructions for use should be followed closely and temperature should be monitored for every test performed. Gathering additional data in summer months to confirm that test specificity remains high with elevated environmental temperatures would be prudent.

Our data are consistent with, or show better performance than, those obtained in other field studies of visually-read Ag RDTs. A recent European prospective study [4] evaluated visually-read Ag RDT test performance at POC versus RT-PCR using nasopharyngeal/oropharyngeal samples and found that the best-performing Ag RDT (Standard Q COVID-19 Ag, SD Biosensor) was 76.6% sensitive and 99.3% specific, with high sensitivity in samples with Ct values < 25, moderate sensitivity in samples with Ct <30, and poor sensitivity above Ct 30 [4]. Another POC study of the SD Q test (NP swab) versus RT-PCR (NP/OP swab) in symptomatic adults at a drive-through site in the Netherlands [12] yielded Ag RDT sensitivity of 84.9% and specificity of 99.5%, with 95.8% sensitivity in people within 7 days of symptoms and with Ct <30. A recent study of the Abbott BinaxNOW versus AN swab RT-PCR as performed in adults on a public plaza in San Francisco found that the test had 93.3% sensitivity in samples with Ct <30, and 99.9% specificity [9]. However, achieving this specificity required an off-label reading procedure in which the reader was asked to disregard bands that did not extend across the full width of the strip. We did observe occasional inconsistent band intensity across the width of some test bands, but did not identify a correlation between that finding and false positivity. All twelve false positive tests in our study had faint positive (and otherwise normal) bands scored by both independent readers. The San Francisco study also used two readers for each test, with a tie breaker read if they disagreed, but did not report data on inter-operator agreement. A study of the Abbott PanBio test in symptomatic adults and children in Spain [11] compared POC Ag RDT (NP swab) to RT-PCR (NP swab in 3.0 mL media) and found overall Ag RDT sensitivity of 79.6%, with specificity of 100%. Notably, sensitivity was higher in adults (82.6%) than children (62.5%), and sensitivity in individuals with RT-PCR Ct <25 (viral load >5.9 log10 copies/mL) was 100%; our own observations are consistent with both findings. The authors speculated that the lower sensitivity in symptomatic children versus adults might be due to more difficulty in pinpointing date of symptom onset in children, which is also a possibility in our study.

However, an alternative explanation is that symptomatic children have a different distribution of viral loads than symptomatic adults, with a higher proportion having lower viral loads (e.g. above Ct of 30) within the first week of symptoms. This hypothesis is supported by distributions of Ct values (and antigen concentrations) in NP swab samples from symptomatic adults versus children in a recent study [16], but other studies of this question have yielded mixed results [19-21]. Defining viral load distributions and kinetics in symptomatic children remains an important area for research.

Key considerations for use of Ag RDTs are whether they are able to reliably identify patients with high loads of live virus that may be more at risk for transmitting disease, and whether infected individuals missed by Ag RDTs might or might not be infectious to others. Several reports have noted that virus could only rarely be cultured from patient samples with measured viral loads below approximately 1×10^5^ RNA copies/mL [19, 22-24]. Notably, Albert et al. found that SARS-CoV-2 could not be cultured from specimens from 11 individuals with positive RT-PCR/negative POC Abbott PanBio Ag RDT results, all with <5.9 log10 copies/mL [11], and Igloi et al found that most culture-positive individuals were also positive by the SD Q assay [12].

Overlap between viral culture and Ag RDT percent positivity (versus RT-PCR) has also been described for the BD Veritor System [25]. However, virus has been recovered from samples with RNA levels as low as 1.2×10^4^ copies/mL [26], and from samples with a Ct value of 34-35 on a range of RT-PCR assays [27-29]. Multiple evaluations of POC Ag RDT performance have assessed performance at Ct value cutoffs of 25 and 30 (e.g. [4, 9]), which guided our choice to assess sensitivity of the BinaxNOW at three different Ct cutoffs: 25, 30, and 35. For the RT-PCR assay used in this study, Ct values of 25, 30, and 35 correspond to approximately 5.4×10^5^, 1.7×10^4^, and 5.5×10^2^ copies/mL, respectively (Niall Lennon, personal communication), and we found that patients with Ct values ≤30 were reliably detected (95.8%) by the BinaxNOW, similar to results from Pilarowski et al (93.3% sensitivity with Ct <30, corresponding to 1.9 x 10^4^ copies/mL [9]). A recent study of multiple commercial Ag RDTs found that the analytical sensitivity range overlapped with viral load ranges typically observed in the first week of symptoms, also considered to be the period of highest infectiousness [18]; consistent with this, we observed that the sensitivity of the BinaxNOW in adults with ≤7 days of symptoms was 96.5%.

Our study had some limitations. We recognize that the comparator in our study was RT-PCR performed on an AN swab, as opposed to an NP swab, which is still considered the reference method by the FDA [30]. While AN swabs have had lower sensitivity than NP swabs in some studies, the sensitivity is highly dependent on the sampling technique and assay used [31]. The dry AN swab sampling method used in this study has been shown to have similar sensitivity to paired NP swabs in transport media [17]. We also note that a recent comparison study demonstrated that Ag RDT performance with AN swabs was similar to Ag RDT performance with NP swabs [5]. We were not able to have all tests read by two independent readers, but did so for the majority of tests. Finally, we recognize that our symptomatic pediatric cohort was relatively small and thus the confidence interval relatively wide, exemplifying the challenges in assessing COVID-19 diagnostics in children. There may be important differences in performance characteristics of Ag tests or in viral dynamics corresponding to ages of children, but there were insufficient symptomatic children with positive tests in the study to stratify by age group.

Current recommendations from the WHO [32] suggest that Ag RDTs be used in symptomatic individuals within the first 5-7 days of symptoms, with repeat testing or RT-PCR confirmation of negative Ag RDT results if possible. WHO recommendations include testing during suspected outbreaks, monitoring trends in disease incidence, and screening for early detection/isolation when widespread community transmission exists, but operationalizing testing under these scenarios is complicated by the recommendations for confirmation by RT-PCR of both positive (outbreak) and negative (symptomatic patient) results. Centers for Disease Control and Prevention recommendations for use of antigen tests were recently updated and address use of antigen tests (with/without NAAT confirmation) in various testing scenarios based on data to date [33].

Our study data provides the following practical considerations for use of the BinaxNOW test in different populations, taking into account current conditions in MA as reflected in prevalence at this testing site (asymptomatic >5-10%; symptomatic >20%):

The test had particularly strong performance in adults with symptoms for ≤ 7 days, with very high sensitivity (96.5%), specificity (100%), PPV (100%), and NPV (98.9%). In children with symptoms ≤7 days, specificity (100%) and PPV (100%) were very high, but sensitivity was lower than in adults (84.6% versus 96.5%) though still acceptable per the FDA’s target of ≥ 80% [34, 35]); NPV was 94.2%. Collection of additional data in symptomatic children will help further support these conclusions.

In asymptomatic individuals, the BinaxNOW test had very high specificity in both children (99.0%) and adults (99.6%); PPV was the same in children (90.9%) and adults (90.9%). Sensitivity in children (65.4%) and adults (70.2%) was low. Thus, the test does not appear to be optimal for ruling out SARS-CoV-2 infection in asymptomatic adults or children; use in serial testing programs and for testing of contacts of known cases deserves independent study. FDA does provide guidance for consideration of serial antigen testing if the sensitivity is lower, e.g. 70% [34, 35]. It should be noted that in all groups, BinaxNOW sensitivity followed Ct value distribution, with 95.8% sensitivity observed in all individuals with Ct ≤ 30. Therefore, false negative BinaxNOW results were largely confined to those perhaps least likely to transmit SARS-CoV-2.

Sensitivity (66.7%) was low in adults with symptoms for > 7 days, and there were too few children with symptoms > 7 days to draw conclusions about performance. The test is not currently recommended for use in individuals with symptoms > 7 days.

Quality Control (QC) testing should be performed per manufacturer’s recommendations, with weekly repeat of QC samples if the kit has not been fully used. It would be prudent to reevaluate test specificity in warm conditions; careful attention to temperature is critical. A skilled laboratorian can perform and read 20 tests per hour. Excellent inter-operator agreement indicates that an individual can perform and read the BinaxNOW test alone.

## Data Availability

All data referred to in the manuscript are publicly available.

## Acknowledgments

We thank Kerin Milesky (Director, Office of Emergency Preparedness, Massachusetts Department of Public Health), Samantha Phillips (Director, Massachusetts Emergency Management Agency), and members of the Massachusetts State Fire Services (Bill Pappas, Mike Burnell, Emit Magdeleno, Greg Jackson, Ed Loader) for assistance with logistical and support assistance at the Lawrence General Hospital Stop the Spread site. We acknowledge the assistance of the members of the Epidemiology Division of the Bureau of Infectious Disease and Laboratory Sciences of the Massachusetts Department of Public Health.

This publication was supported by Cooperative Agreement Number 1U60OE000103, funded by Centers for Disease Control and Prevention through the Association of Public Health Laboratories. The findings and conclusions in this report are those of the authors and do not necessarily represent the official position of the Centers for Disease Control and Prevention (CDC) nor the official views of the Association of Public Health Laboratories.

