## Supplementary Materials for "Performance and Implementation Evaluation of the Abbott BinaxNOW Rapid Antigen Test in a High-throughput Drive-through Community Testing Site in Massachusetts"

**Supplementary Methods**

Anterior Nasal Swab Collection procedure

The following swab collection procedure was utilized: after the participant blew their nose, the collector inserted swab #1 into the individual's right nostril and rotated the swab 5 times in a circular motion around the inside wall of the nostril for a duration of 10-15 seconds, followed by a repeat of the process in the left nostril. The swab was placed into a sterile plastic tube. Immediately thereafter, swab #2 was inserted into the individual's left nostril (always opposite from the initial swab) and the process repeated (left nostril followed by right nostril). To minimize sampling bias, the study site personnel alternated which test-specific nasal swab sample was collected first (BinaxNOW or RT-PCR).

Kit Storage

Abbott BinaxNOW kits were stored in a temperature-controlled office (70-72°F) and transferred to the testing site as needed.

Testing Site

Testing was performed in a "testing pod," initially a shipping container. The pod was initially open on one side to the outdoors, but difficulty in maintaining adequate temperature in the pod required moving testing to a fully enclosed room (mobile trailer). The testing site contained three 6' tables. Two tables were designated as "dirty" for testing purposes and one "clean" for data entry. Absorbent underpads with plastic backing (Attends ASB-2336, Greenville, NC) were placed on testing tables along with hand sanitizer, gloves, testing kits, pens, sticky notes, a continuous temperature and humidity data logger (HOBO UX100-011A, Bourne, MA), a digital clock, and a bin for specimens to be tested. The clean table included hand sanitizer, gloves, pens, a time stamp machine, and a laptop computer for data entry. Underpads were removed daily, surfaces were disinfected with germicidal disposable wipes (PDI, Woodcliff Lake, NJ), and new underpads placed. Operators performing testing were wearing disposable gowns, N95 respirators, face shields and gloves.

Quality Control

Lot numbers were recorded for Abbott BinaxNOW kits (boxes of 40 tests), test cards, manufacturer-provided swabs [unflocked foam tipped; Puritan #25-1506 (Guilford, ME)], positive controls, and extraction reagent. Manufacture and expiration dates were also recorded. Each study subject tested was recorded for every kit of BinaxNow tests used. Positive and negative controls were run for every kit when first opened. If an opened kit was not used in full over the course of one day, a negative control was run for the kit the following day when testing resumed.

BinaxNOW Testing

At the collection site, anterior nasal swab specimens were collected using the manufacturer-provided swab (above) into a labeled 15ml conical tube and placed into a specimen collection bag with a BinaxNow intake form (which was labeled and time/date stamped at the time of collection) and extra labels. Specimens were then shuttled to the testing pod by a designated individual ("runner"), either on foot (shipping container) or by car (mobile trailer). All tests were initiated within an hour of specimen collection time. For each specimen, testing staff added 6 drops of extraction reagent to the BinaxNOW card (making sure to remain perpendicular and avoid touching the tip of the reagent bottle to the card), inserted the swab specimen and rotated three times, closed the testing card, and affixed a patient label to the outside of the BinaxNOW card. The time the card was closed was recorded on the BinaxNOW intake form as the test start time. The test start time and time the card was to be read (16 minutes after closing the card) were also recorded on a sticky note and adhered to the outside of the BinaxNOW card. At the time of result reading, the tester silently recorded their result, initials, and time on the Binax intake form as the official result. The tester then passed the card to a second reader, who read the card result aloud (so that the tester could record results on the back of the intake form) and took a photo of the card. There was no attempt to resolve any discordance between the two readers; the first read of each card was the official result used for database and clinical reporting. If the result was determined to be positive by either reader, the reader categorized the positive line as “faint”, “medium”, or “strong” depending on the strength of the positive sample line compared to the control line of each card. Completed BinaxNOW cards were discarded in biohazard waste.

PCR assay details

Dry swabs (Fisher Scientific, # NC1817884) transported in a sterile tube (Becton Dickinson, # BD 366408) at room temperature were resuspended in 1000 uL of Swab Preservation Buffer (Norgen Biotek, Inc) and allowed to elute for 15 minutes. Extraction and RT-PCR methods followed the EUA protocol for the CRSP SARS-CoV-2 Real-time Reverse Transcriptase (RT)-PCR Diagnostic Assay (https://www.fda.gov/media/139858/download); the target is the N2 gene with a cycle threshold cut off value of 40.

### Supplementary Table 1
### Abbott BinaxNow results, RT-PCR Ct values, and symptoms in RT-PCR-positive adults

| Age | Sex | PCR Result | Ct (N2) | Ct (RP) | Binax Result | COVID-19 symptoms? | Sore throat? | Cough? | Chills? | Body Aches? | Short of Breath? | Fever? | Runny Nose? | Congestion? | Nausea? | Vomiting? | Diarrhea? | Loss of Taste or Smell? | # Days Since Symptom Onset |
|---|---|---|---|---|---|---|---|---|---|---|---|---|---|---|---|---|---|---|---|
| 21 | F | POS | 34.26 | 19.58 | NEG | ASx | | | | | | | | | | | | | n/a |
| 23 | M | POS | 35.48 | 25.16 | NEG | ASx | | | | | | | | | | | | | n/a |
| 26 | M | POS | 35.43 | 25.88 | NEG | ASx | | | | | | | | | | | | | n/a |
| 27 | M | POS | 35.17 | 24.26 | NEG | ASx | | | | | | | | | | | | | n/a |
| 28 | F | POS | 34.66 | 26.29 | NEG | ASx | | | | | | | | | | | | | n/a |
| 30 | M | POS | 35.19 | 20.77 | NEG | ASx | | | | | | | | | | | | | n/a |
| 33 | M | POS | 32.76 | 24.52 | NEG | ASx | | | | | | | | | | | | | n/a |
| 34 | M | POS | 32.36 | 22.42 | NEG | ASx | | | | | | | | | | | | | n/a |
| 38 | F | POS | 31.22 | 21.78 | NEG | ASx | | | | | | | | | | | | | n/a |
| 39 | M | POS | 37.03 | 28.61 | NEG | ASx | | | | | | | | | | | | | n/a |
| 48 | M | POS | 35.29 | 27.46 | NEG | ASx | | | | | | | | | | | | | n/a |
| 50 | F | POS | 34.87 | 21.31 | NEG | ASx | | | | | | | | | | | | | n/a |
| 50 | F | POS | 34.73 | 24.63 | NEG | ASx | | | | | | | | | | | | | n/a |
| 56 | F | POS | 32.3 | 26.75 | NEG | ASx | | | | | | | | | | | | | n/a |
| 60 | F | POS | 31.6 | 26.5 | NEG | ASx | | | | | | | | | | | | | n/a |
| 71 | M | POS | 33.2 | 19.62 | NEG | ASx | | | | | | | | | | | | | n/a |
| 91 | M | POS | 34.56 | 24.54 | NEG | ASx | | | | | | | | | | | | | n/a |
| 21 | M | POS | 14.62 | 23.23 | POS | ASx | | | | | | | | | | | | | n/a |
| 22 | F | POS | 32.89 | 30.74 | POS | ASx | | | | | | | | | | | | | n/a |
| 22 | M | POS | 27.74 | 22.13 | POS | ASx | | | | | | | | | | | | | n/a |
| 23 | F | POS | 18.75 | 25.23 | POS | ASx | | | | | | | | | | | | | n/a |
| 23 | F | POS | 26.75 | 24.18 | POS | ASx | | | | | | | | | | | | | n/a |
| 24 | M | POS | 29.67 | 21.5 | POS | ASx | | | | | | | | | | | | | n/a |
| 25 | M | POS | 14.1 | 19.83 | POS | ASx | | | | | | | | | | | | | n/a |
| 27 | F | POS | 20.69 | 27.21 | POS | ASx | | | | | | | | | | | | | n/a |
| 28 | M | POS | 23.63 | 27.99 | POS | ASx | | | | | | | | | | | | | n/a |
| 29 | F | POS | 22.21 | 24.08 | POS | ASx | | | | | | | | | | | | | n/a |
| 31 | F | POS | 17.55 | 27.16 | POS | ASx | | | | | | | | | | | | | n/a |
| 31 | M | POS | 14.57 | 22.16 | POS | ASx | | | | | | | | | | | | | n/a |
| 32 | M | POS | 28.17 | 24.67 | POS | ASx | | | | | | | | | | | | | n/a |
| 34 | F | POS | 22.47 | 27.53 | POS | ASx | | | | | | | | | | | | | n/a |
| 35 | F | POS | 17.39 | 23.93 | POS | ASx | | | | | | | | | | | | | n/a |
| 38 | M | POS | 26.95 | 24.65 | POS | ASx | | | | | | | | | | | | | n/a |
| 39 | F | POS | 20.82 | 29.82 | POS | ASx | | | | | | | | | | | | | n/a |
| 42 | F | POS | 13.65 | 22.24 | POS | ASx | | | | | | | | | | | | | n/a |
| 42 | F | POS | 28.4 | 25.58 | POS | ASx | | | | | | | | | | | | | n/a |
| 46 | M | POS | 12.82 | 20.87 | POS | ASx | | | | | | | | | | | | | n/a |
| 48 | F | POS | 16.4 | 24.47 | POS | ASx | | | | | | | | | | | | | n/a |
| 49 | F | POS | 22.3 | 23.42 | POS | ASx | | | | | | | | | | | | | n/a |
| 50 | M | POS | 16.35 | 24.14 | POS | ASx | | | | | | | | | | | | | n/a |
| 51 | M | POS | 26.91 | 23.37 | POS | ASx | | | | | | | | | | | | | n/a |
| 51 | M | POS | 17.16 | 23.86 | POS | ASx | | | | | | | | | | | | | n/a |
| 53 | F | POS | 25.78 | 23.3 | POS | ASx | | | | | | | | | | | | | n/a |
| 53 | F | POS | 21.4 | 23.19 | POS | ASx | | | | | | | | | | | | | n/a |
| 54 | F | POS | 28.3 | 21.6 | POS | ASx | | | | | | | | | | | | | n/a |
| 57 | F | POS | 21.66 | 24.84 | POS | ASx | | | | | | | | | | | | | n/a |
| 60 | M | POS | 28.65 | 26.5 | POS | ASx | | | | | | | | | | | | | n/a |
| 61 | F | POS | 15.64 | 26.47 | POS | ASx | | | | | | | | | | | | | n/a |
| 61 | M | POS | 24.4 | 27.67 | POS | ASx | | | | | | | | | | | | | n/a |
| 61 | M | POS | 21.61 | 27.39 | POS | ASx | | | | | | | | | | | | | n/a |
| 63 | F | POS | 25.94 | 28.19 | POS | ASx | | | | | | | | | | | | | n/a |
| 64 | M | POS | 32.01 | 23.85 | POS | ASx | | | | | | | | | | | | | n/a |
| 65 | M | POS | 26.09 | 21.92 | POS | ASx | | | | | | | | | | | | | n/a |
| 67 | F | POS | 15.4 | 24.6 | POS | ASx | | | | | | | | | | | | | n/a |
| 71 | F | POS | 28.45 | 22.76 | POS | ASx | | | | | | | | | | | | | n/a |
| 71 | M | POS | 32.83 | 25.58 | POS | ASx | | | | | | | | | | | | | n/a |
| 79 | F | POS | 20.55 | 24.67 | POS | ASx | | | | | | | | | | | | | n/a |
| 29 | M | POS | 32.54 | 24.93 | NEG | Sx | | Y | Y | | | Y | Y | Y | | | | | 2 |
| 32 | F | POS | 31.56 | 20.47 | NEG | Sx | | Y | Y | Y | | | Y | Y | | | | | 2 |
| 63 | M | POS | 33.77 | 24.45 | NEG | Sx | | Y | | | | | | | | | | | 1 |
| 19 | M | POS | 26.23 | 23.73 | POS | Sx | Y | | Y | Y | | | | | | | | | 3 |
| 20 | F | POS | 20.47 | 22.8 | POS | Sx | Y | Y | Y | Y | | | Y | Y | | | | | 3 |
| 20 | M | POS | 27.87 | 24.95 | POS | Sx | Y | Y | | | | | Y | Y | | | | | 4 |
| 20 | M | POS | 14.55 | 22.4 | POS | Sx | | | | | | | Y | | | | | | 0 |
| 21 | F | POS | 17.94 | 23.13 | POS | Sx | | Y | Y | | Y | | Y | Y | | | | | 2 |
| 21 | M | POS | 16.28 | 23.96 | POS | Sx | | Y | Y | Y | | Y | Y | Y | | | | | 2 |
| 22 | F | POS | 20.59 | 21.94 | POS | Sx | | Y | | | Y | | Y | Y | | | Y | Y | 3 |
| 23 | F | POS | 20.16 | 27.91 | POS | Sx | | | | Y | | | Y | Y | | Y | | Y | 3 |
| 23 | F | POS | 27.72 | 28.5 | POS | Sx | Y | Y | Y | Y | | Y | | | Y | | Y | | 2 |
| 24 | M | POS | 20.41 | 28.39 | POS | Sx | Y | | Y | Y | | Y | Y | Y | | | | Y | 1 |
| 25 | M | POS | 29.26 | 22.36 | POS | Sx | Y | Y | | Y | | | Y | Y | | | | Y | 4 |
| 25 | F | POS | 20.81 | 25.63 | POS | Sx | | Y | | | | Y | Y | | | | | Y | 2 |
| 25 | F | POS | 19.47 | 21.79 | POS | Sx | Y | Y | | Y | | | Y | Y | | | | Y | 3 |
| 25 | M | POS | 26.26 | 24.39 | POS | Sx | Y | Y | Y | Y | Y | Y | Y | | | | | | 3 |
| 26 | M | POS | 15.62 | 24.26 | POS | Sx | Y | | | | | | Y | Y | | | | | 1 |
| 26 | F | POS | 19.19 | 22.21 | POS | Sx | Y | Y | | | | | | | | | | | 2 |
| 27 | F | POS | 17.79 | 25.23 | POS | Sx | Y | Y | Y | Y | | | Y | Y | | | | | 2 |

| | | | | | | | | | | | | | | | | | | | |
|---|---|---|---|---|---|---|---|---|---|---|---|---|---|---|---|---|---|---|---|
| 27 | M | POS | 15.95 | 24.3 | POS | Sx | Y | | | Y | | | Y | Y | Y | | | | 2 |
| 27 | F | POS | 18.81 | 23.78 | POS | Sx | | Y | Y | Y | | Y | Y | | | | | | 2 |
| 28 | M | POS | 25.41 | 24.61 | POS | Sx | | | Y | Y | | Y | | Y | | Y | Y | | 4 |
| 29 | F | POS | 16.5 | 21.78 | POS | Sx | | | | | | | | | | | | Y | 0 |
| 29 | M | POS | 18.2 | 23.42 | POS | Sx | | | | | | | Y | | | | | | 1 |
| 30 | M | POS | 27.23 | 29.61 | POS | Sx | | Y | Y | Y | | | Y | Y | Y | | | | 2 |
| 31 | F | POS | 25.48 | 24.96 | POS | Sx | Y | Y | Y | Y | Y | Y | Y | | | | | | 3 |
| 31 | F | POS | 18.33 | 25.67 | POS | Sx | | Y | Y | | | | | Y | | | | | 1 |
| 31 | F | POS | 21.89 | 26.98 | POS | Sx | | Y | | Y | Y | | Y | Y | Y | | | Y | 4 |
| 31 | F | POS | 15.21 | 23.27 | POS | Sx | Y | Y | Y | Y | Y | Y | Y | Y | Y | Y | Y | Y | 4 |
| 31 | M | POS | 16.17 | 21.38 | POS | Sx | | Y | Y | Y | | Y | Y | Y | Y | | Y | Y | 0 |
| 32 | F | POS | 29.47 | 26.43 | POS | Sx | | | | Y | | Y | Y | Y | | Y | | | 6 |
| 32 | F | POS | 13.77 | 24.14 | POS | Sx | | | | | | | Y | | | | | | 1 |
| 32 | M | POS | 26.76 | 24.76 | POS | Sx | | Y | | Y | | Y | | Y | | | | Y | 3 |
| 33 | M | POS | 24.36 | 21.6 | POS | Sx | | | Y | | | | | Y | | | | Y | 3 |
| 33 | F | POS | 14.32 | 22.84 | POS | Sx | Y | Y | | Y | | | Y | | | | | | 1 |
| 33 | F | POS | 17.12 | 21.83 | POS | Sx | | Y | | Y | | | Y | Y | | | | | 1 |
| 34 | M | POS | 27.63 | 26.65 | POS | Sx | | Y | | Y | | | | Y | | | Y | | 2 |
| 34 | F | POS | 30.52 | 23.25 | POS | Sx | Y | | Y | | | Y | Y | | | Y | Y | | 5 |
| 34 | M | POS | 18.75 | 22.42 | POS | Sx | Y | Y | | Y | | | Y | | | | | | 3 |
| 35 | F | POS | 22.88 | 28.59 | POS | Sx | | Y | Y | Y | | | | | | Y | | Y | 1 |
| 35 | F | POS | 16.4 | 22.27 | POS | Sx | Y | | | Y | | | | | | | | | 1 |
| 35 | F | POS | 20.19 | 22.43 | POS | Sx | Y | Y | Y | Y | | | Y | Y | | | Y | Y | 3 |
| 35 | M | POS | 25.06 | 26.91 | POS | Sx | Y | Y | | | | | Y | | | | | Y | 2 |
| 35 | M | POS | 27.25 | 22.99 | POS | Sx | Y | Y | Y | | | | Y | Y | Y | | | | 3 |
| 36 | F | POS | 22.81 | 27.54 | POS | Sx | | Y | | Y | | | Y | | | | | | 1 |
| 37 | F | POS | 19.5 | 26.99 | POS | Sx | Y | | Y | Y | | Y | Y | Y | | | Y | | 2 |
| 37 | M | POS | 22.18 | 21.61 | POS | Sx | Y | Y | Y | | | | | | | | | | 2 |
| 38 | M | POS | 18.33 | 27.29 | POS | Sx | Y | Y | Y | | Y | Y | Y | | | | | | 6 |
| 38 | F | POS | 31.41 | 22.64 | POS | Sx | Y | Y | Y | Y | Y | | Y | Y | Y | | | | 1 |
| 38 | F | POS | 28.04 | 22.86 | POS | Sx | | | | | Y | | | | | | | Y | 2 |
| 39 | M | POS | 29.14 | 29.32 | POS | Sx | | Y | Y | Y | | Y | Y | Y | | | | Y | 6 |
| 39 | F | POS | 15.41 | 22.8 | POS | Sx | Y | Y | Y | Y | Y | Y | Y | | | | Y | | 1 |
| 40 | M | POS | 20.77 | 24.58 | POS | Sx | | Y | Y | Y | | Y | | | | | | Y | 1 |
| 40 | F | POS | 21.26 | 25.98 | POS | Sx | Y | Y | Y | Y | | Y | Y | Y | | | | | 5 |
| 40 | M | POS | 23.03 | 20.72 | POS | Sx | Y | | | | | | Y | | | | | Y | 2 |
| 41 | M | POS | 17.23 | 24.19 | POS | Sx | | | | | Y | | | | | | | | 3 |
| 41 | M | POS | 19.24 | 21.43 | POS | Sx | Y | Y | | Y | Y | Y | | | | | | | 2 |
| 42 | M | POS | 23.2 | 24.16 | POS | Sx | Y | Y | Y | Y | Y | | Y | Y | | | Y | | 2 |
| 44 | F | POS | 17.67 | 25.61 | POS | Sx | | Y | Y | | | | | | | | | | 1 |
| 44 | F | POS | 15.2 | 22.19 | POS | Sx | Y | Y | | Y | Y | | | | | | | | 3 |
| 45 | M | POS | 18.7 | 23.76 | POS | Sx | | Y | Y | | | | | | | | | | 3 |
| 45 | F | POS | 21.15 | 24.76 | POS | Sx | Y | Y | Y | Y | | Y | Y | Y | | | | | 3 |
| 46 | F | POS | 15.2 | 27.54 | POS | Sx | Y | Y | | Y | | | Y | | | | | | 2 |
| 47 | F | POS | 32.6 | 27.59 | POS | Sx | Y | Y | Y | Y | Y | | | Y | Y | | Y | Y | 6 |
| 47 | M | POS | 26.77 | 23.56 | POS | Sx | Y | Y | Y | Y | | Y | | | | | | | 5 |
| 48 | F | POS | 34.58 | 24.96 | POS | Sx | Y | Y | | Y | Y | | Y | Y | | | Y | | 3 |
| 48 | M | POS | 21.82 | 24.66 | POS | Sx | | | Y | Y | | Y | | Y | | | | | 2 |
| 49 | M | POS | 30.73 | 21.95 | POS | Sx | | Y | | | | | | | | | | | 5 |
| 49 | M | POS | 19.16 | 21.67 | POS | Sx | | | | | | Y | | | | | | | 0 |
| 50 | F | POS | 20.45 | 21.07 | POS | Sx | Y | Y | Y | Y | Y | | Y | Y | | | | | 0 |
| 51 | F | POS | 16.57 | 26.99 | POS | Sx | | Y | Y | Y | | Y | Y | | Y | | Y | | 1 |
| 51 | M | POS | 15.46 | 21.47 | POS | Sx | Y | Y | Y | Y | | Y | | | | | | | 1 |
| 54 | M | POS | 24.18 | 24.5 | POS | Sx | | Y | | Y | | Y | | | | | | | 1 |
| 54 | F | POS | 16.92 | 22.82 | POS | Sx | Y | Y | Y | | | Y | | | | | | Y | 3 |
| 54 | M | POS | 11.78 | 18.96 | POS | Sx | | | | Y | Y | Y | | Y | | | | Y | 1 |
| 57 | M | POS | 18.69 | 28.26 | POS | Sx | | | Y | Y | Y | | Y | Y | | | | | 1 |
| 57 | F | POS | 21.8 | 24.73 | POS | Sx | Y | Y | | Y | | Y | | | | | | | 1 |
| 60 | F | POS | 16.18 | 25.74 | POS | Sx | Y | | | | Y | | | | | | | | 1 |
| 61 | M | POS | 18.82 | 25.48 | POS | Sx | | Y | Y | | | Y | Y | | | | | | 2 |
| 65 | F | POS | 14.61 | 23.24 | POS | Sx | | | Y | Y | | | | Y | Y | | | | 3 |
| 66 | M | POS | 15.35 | 18.78 | POS | Sx | Y | Y | Y | Y | Y | Y | Y | Y | Y | Y | | | 3 |
| 67 | M | POS | 33.69 | 25.16 | POS | Sx | | | | | Y | | Y | Y | | | | | 2 |
| 70 | M | POS | 26.78 | 26.3 | POS | Sx | Y | Y | Y | Y | Y | Y | Y | | | | | | 0 |
| 70 | M | POS | 22 | 25.3 | POS | Sx | | Y | | | | | | | | | | | 2 |
| 32 | M | POS | 34.2 | 26.37 | NEG | Sx7+ | | | | Y | Y | | Y | | | | Y | Y | 7+ |
| 32 | F | POS | 34.05 | 23.27 | NEG | Sx7+ | | | | | | | | | | | | Y | 7+ |
| 33 | F | POS | 33.79 | 25.58 | NEG | Sx7+ | | Y | | Y | Y | | | Y | | | Y | | 7+ |
| 43 | M | POS | 34.56 | 25.89 | NEG | Sx7+ | | Y | Y | Y | | | Y | Y | | | | | 7+ |
| 67 | F | POS | 30.4 | 25.95 | NEG | Sx7+ | | | | | | | | Y | | | | | 7+ |
| 19 | F | POS | 22.78 | 22.71 | POS | Sx7+ | | | | | | | Y | Y | | | | | 7+ |
| 20 | M | POS | 29.39 | 22.48 | POS | Sx7+ | | Y | | Y | Y | | Y | Y | | | | | 7+ |
| 25 | F | POS | 29.8 | 24.6 | POS | Sx7+ | | Y | | | | | Y | Y | | | | | 7+ |
| 30 | F | POS | 17.71 | 27.39 | POS | Sx7+ | Y | Y | | | | | | Y | | | | | 7+ |
| 30 | M | POS | 14.71 | 23.66 | POS | Sx7+ | Y | Y | Y | | | | | | | | | | 7+ |
| 34 | M | POS | 25.94 | 26.03 | POS | Sx7+ | Y | Y | | Y | | | Y | | | | | Y | 7+ |
| 43 | F | POS | 27.12 | 19.54 | POS | Sx7+ | | | | | | | | | | | | Y | 7+ |
| 50 | F | POS | 34.56 | 24.45 | POS | Sx7+ | | | | | | | Y | Y | | | | Y | 7+ |
| 53 | M | POS | 17.8 | 23.87 | POS | Sx7+ | | Y | Y | | Y | | | | Y | | | | 7+ |
| 68 | M | POS | 20.24 | 23.53 | POS | Sx7+ | | Y | | | Y | | | | | | Y | | 7+ |

F, female; M, male; POS, positive; Ct, cycle threshold; N2, SARS-CoV-2 target; RP, host control target; ASx, asymptomatic; Sx, symptomatic; n/a, not applicable

### Supplementary Table 2

### Abbott BinaxNow results, RT-PCR Ct values, and symptoms in RT-PCR-positive children

| Age | Sex | PCR Result | Ct (N2) | Ct (RP) | Binax Result | COVID-19 symptoms? | Sore throat? | Cough? | Chills? | Body Aches? | Short of Breath? | Fever? | Runny Nose? | Congestion? | Nausea? | Vomiting? | Diarrhea? | Loss of Taste or Smell? | # Days Since Symptom Onset |
|---|---|---|---|---|---|---|---|---|---|---|---|---|---|---|---|---|---|---|---|
| 0 | F | POS | 33.59 | 21.55 | NEG | ASx | | | | | | | | | | | | | n/a |
| 0 | M | POS | 29.89 | 20.61 | NEG | ASx | | | | | | | | | | | | | n/a |
| 1 | M | POS | 34.26 | 21.97 | NEG | ASx | | | | | | | | | | | | | n/a |
| 2 | M | POS | 37.1 | 26.4 | NEG | ASx | | | | | | | | | | | | | n/a |
| 3 | F | POS | 31.7 | 21.76 | NEG | ASx | | | | | | | | | | | | | n/a |
| 3 | F | POS | 26.7 | 19.84 | NEG | ASx | | | | | | | | | | | | | n/a |
| 3 | F | POS | 36.51 | 24.03 | NEG | ASx | | | | | | | | | | | | | n/a |
| 3 | M | POS | 28.93 | 21.76 | NEG | ASx | | | | | | | | | | | | | n/a |
| 3 | M | POS | 32.44 | 24.73 | NEG | ASx | | | | | | | | | | | | | n/a |
| 3 | F | POS | 32.31 | 22.46 | NEG | ASx | | | | | | | | | | | | | n/a |
| 5 | M | POS | 32.87 | 26.98 | NEG | ASx | | | | | | | | | | | | | n/a |
| 6 | F | POS | 26.82 | 22.58 | NEG | ASx | | | | | | | | | | | | | n/a |
| 7 | F | POS | 33.01 | 20.2 | NEG | ASx | | | | | | | | | | | | | n/a |
| 7 | F | POS | 30.99 | 21.65 | NEG | ASx | | | | | | | | | | | | | n/a |
| 7 | F | POS | 33.72 | 23.35 | NEG | ASx | | | | | | | | | | | | | n/a |
| 7 | M | POS | 36.31 | 24.21 | NEG | ASx | | | | | | | | | | | | | n/a |
| 8 | M | POS | 34.62 | 22.23 | NEG | ASx | | | | | | | | | | | | | n/a |
| 9 | M | POS | 34.34 | 22.58 | NEG | ASx | | | | | | | | | | | | | n/a |
| 9 | M | POS | 27.42 | 21.64 | NEG | ASx | | | | | | | | | | | | | n/a |
| 10 | F | POS | 32.47 | 22.45 | NEG | ASx | | | | | | | | | | | | | n/a |
| 10 | M | POS | 34.07 | 25.45 | NEG | ASx | | | | | | | | | | | | | n/a |
| 12 | F | POS | 27.45 | 24.01 | NEG | ASx | | | | | | | | | | | | | n/a |
| 12 | M | POS | 39.84 | 24.35 | NEG | ASx | | | | | | | | | | | | | n/a |
| 12 | M | POS | 23.43 | 20.22 | NEG | ASx | | | | | | | | | | | | | n/a |
| 12 | F | POS | 32.83 | 24.29 | NEG | ASx | | | | | | | | | | | | | n/a |
| 13 | F | POS | 26.89 | 21.82 | NEG | ASx | | | | | | | | | | | | | n/a |
| 14 | F | POS | 32.78 | 23.33 | NEG | ASx | | | | | | | | | | | | | n/a |
| 15 | F | POS | 36.84 | 20.19 | NEG | ASx | | | | | | | | | | | | | n/a |
| 15 | F | POS | 33.39 | 22.53 | NEG | ASx | | | | | | | | | | | | | n/a |
| 16 | M | POS | 36.5 | 19.31 | NEG | ASx | | | | | | | | | | | | | n/a |
| 16 | F | POS | 33.08 | 22.33 | NEG | ASx | | | | | | | | | | | | | n/a |
| 16 | F | POS | 33.5 | 19.98 | NEG | ASx | | | | | | | | | | | | | n/a |
| 16 | F | POS | 35.4 | 22.32 | NEG | ASx | | | | | | | | | | | | | n/a |
| 17 | F | POS | 34.88 | 24.47 | NEG | ASx | | | | | | | | | | | | | n/a |
| 17 | M | POS | 34.47 | 25.58 | NEG | ASx | | | | | | | | | | | | | n/a |
| 18 | F | POS | 32.22 | 21.37 | NEG | ASx | | | | | | | | | | | | | n/a |
| 18 | F | POS | 33.9 | 26.17 | NEG | ASx | | | | | | | | | | | | | n/a |
| 0 | M | POS | 13.48 | 23.3 | POS | ASx | | | | | | | | | | | | | n/a |
| 0 | F | POS | 17.72 | 27.64 | POS | ASx | | | | | | | | | | | | | n/a |
| 0 | F | POS | 32.61 | 20.25 | POS | ASx | | | | | | | | | | | | | n/a |
| 1 | F | POS | 27.34 | 21.61 | POS | ASx | | | | | | | | | | | | | n/a |
| 1 | M | POS | 14.35 | 20.71 | POS | ASx | | | | | | | | | | | | | n/a |
| 1 | M | POS | 19.05 | 26.8 | POS | ASx | | | | | | | | | | | | | n/a |
| 2 | F | POS | 29.77 | 25.68 | POS | ASx | | | | | | | | | | | | | n/a |
| 2 | F | POS | 25.61 | 19.76 | POS | ASx | | | | | | | | | | | | | n/a |
| 2 | M | POS | 16.75 | 20.83 | POS | ASx | | | | | | | | | | | | | n/a |
| 2 | F | POS | 21.51 | 20.22 | POS | ASx | | | | | | | | | | | | | n/a |
| 2 | F | POS | 22.88 | 20.41 | POS | ASx | | | | | | | | | | | | | n/a |
| 3 | F | POS | 23.96 | 22.89 | POS | ASx | | | | | | | | | | | | | n/a |
| 3 | F | POS | 20.94 | 20.45 | POS | ASx | | | | | | | | | | | | | n/a |
| 3 | M | POS | 25.56 | 20.35 | POS | ASx | | | | | | | | | | | | | n/a |
| 4 | M | POS | 21.68 | 19 | POS | ASx | | | | | | | | | | | | | n/a |
| 5 | M | POS | 27.84 | 18.48 | POS | ASx | | | | | | | | | | | | | n/a |
| 6 | F | POS | 29.61 | 19.83 | POS | ASx | | | | | | | | | | | | | n/a |
| 6 | M | POS | 17.63 | 21.84 | POS | ASx | | | | | | | | | | | | | n/a |
| 6 | M | POS | 13.11 | 20.96 | POS | ASx | | | | | | | | | | | | | n/a |
| 6 | M | POS | 21.53 | 24.64 | POS | ASx | | | | | | | | | | | | | n/a |
| 7 | M | POS | 27.57 | 22.55 | POS | ASx | | | | | | | | | | | | | n/a |
| 7 | F | POS | 21.51 | 22.87 | POS | ASx | | | | | | | | | | | | | n/a |
| 7 | M | POS | 14.5 | 23.54 | POS | ASx | | | | | | | | | | | | | n/a |
| 7 | M | POS | 22.1 | 28.85 | POS | ASx | | | | | | | | | | | | | n/a |
| 7 | F | POS | 27.82 | 23.9 | POS | ASx | | | | | | | | | | | | | n/a |
| 7 | F | POS | 29.83 | 20.78 | POS | ASx | | | | | | | | | | | | | n/a |
| 7 | F | POS | 18.13 | 24.45 | POS | ASx | | | | | | | | | | | | | n/a |
| 8 | F | POS | 18.38 | 28.23 | POS | Asx | | | | | | | | | | | | | n/a |
| 8 | F | POS | 15.28 | 22.14 | POS | ASx | | | | | | | | | | | | | n/a |
| 8 | F | POS | 14.66 | 20.35 | POS | ASx | | | | | | | | | | | | | n/a |

| | | | | | | | | | | | | | | | | | | | |
|---|---|---|---|---|---|---|---|---|---|---|---|---|---|---|---|---|---|---|---|
| 8 | F | POS | 32.73 | 21.97 | POS | ASx | | | | | | | | | | | | | n/a |
| 8 | M | POS | 22.99 | 22.95 | POS | ASx | | | | | | | | | | | | | n/a |
| 8 | F | POS | 22.4 | 24.28 | POS | ASx | | | | | | | | | | | | | n/a |
| 9 | F | POS | 19.96 | 22.67 | POS | ASx | | | | | | | | | | | | | n/a |
| 9 | F | POS | 15.45 | 21.72 | POS | ASx | | | | | | | | | | | | | n/a |
| 9 | F | POS | 23.99 | 19.95 | POS | ASx | | | | | | | | | | | | | n/a |
| 9 | M | POS | 34.59 | 22.05 | POS | ASx | | | | | | | | | | | | | n/a |
| 9 | M | POS | 20.54 | 23.4 | POS | ASx | | | | | | | | | | | | | n/a |
| 9 | F | POS | 14.54 | 21.84 | POS | ASx | | | | | | | | | | | | | n/a |
| 10 | M | POS | 35.88 | 25.08 | POS | ASx | | | | | | | | | | | | | n/a |
| 10 | F | POS | 26.85 | 24.5 | POS | ASx | | | | | | | | | | | | | n/a |
| 11 | F | POS | 16.98 | 24.86 | POS | ASx | | | | | | | | | | | | | n/a |
| 11 | M | POS | 33 | 26.48 | POS | ASx | | | | | | | | | | | | | n/a |
| 11 | M | POS | 29.63 | 20.77 | POS | ASx | | | | | | | | | | | | | n/a |
| 12 | F | POS | 17.77 | 23.91 | POS | Asx | | | | | | | | | | | | | n/a |
| 12 | F | POS | 19.38 | 21.69 | POS | ASx | | | | | | | | | | | | | n/a |
| 12 | M | POS | 28.13 | 24.42 | POS | ASx | | | | | | | | | | | | | n/a |
| 12 | M | POS | 26.13 | 21.43 | POS | ASx | | | | | | | | | | | | | n/a |
| 13 | F | POS | 21.41 | 26.51 | POS | ASx | | | | | | | | | | | | | n/a |
| 13 | M | POS | 15.02 | 23.09 | POS | ASx | | | | | | | | | | | | | n/a |
| 13 | M | POS | 18.81 | 22.28 | POS | ASx | | | | | | | | | | | | | n/a |
| 14 | M | POS | 23 | 23.72 | POS | ASx | | | | | | | | | | | | | n/a |
| 14 | F | POS | 34.35 | 25.54 | POS | ASx | | | | | | | | | | | | | n/a |
| 14 | F | POS | 25.82 | 24.55 | POS | ASx | | | | | | | | | | | | | n/a |
| 15 | M | POS | 23.74 | 20.75 | POS | ASx | | | | | | | | | | | | | n/a |
| 15 | M | POS | 32.1 | 22.45 | POS | ASx | | | | | | | | | | | | | n/a |
| 15 | F | POS | 21.48 | 22.93 | POS | ASx | | | | | | | | | | | | | n/a |
| 15 | M | POS | 24.18 | 22.59 | POS | ASx | | | | | | | | | | | | | n/a |
| 15 | M | POS | 16.09 | 23.35 | POS | ASx | | | | | | | | | | | | | n/a |
| 15 | F | POS | 21.04 | 23.6 | POS | ASx | | | | | | | | | | | | | n/a |
| 15 | F | POS | 20.9 | 23.6 | POS | ASx | | | | | | | | | | | | | n/a |
| 15 | F | POS | 30.4 | 25.71 | POS | ASx | | | | | | | | | | | | | n/a |
| 16 | M | POS | 16.99 | 21.01 | POS | ASx | | | | | | | | | | | | | n/a |
| 16 | F | POS | 18.29 | 23.6 | POS | ASx | | | | | | | | | | | | | n/a |
| 17 | F | POS | 19.81 | 21.22 | POS | ASx | | | | | | | | | | | | | n/a |
| 17 | F | POS | 18.1 | 22.44 | POS | ASx | | | | | | | | | | | | | n/a |
| 17 | F | POS | 17.2 | 24.71 | POS | ASx | | | | | | | | | | | | | n/a |
| 17 | F | POS | 24.45 | 24.72 | POS | ASx | | | | | | | | | | | | | n/a |
| 17 | F | POS | 21.3 | 26.47 | POS | ASx | | | | | | | | | | | | | n/a |
| 18 | F | POS | 35.73 | 23.96 | POS | ASx | | | | | | | | | | | | | n/a |
| 12 | F | POS | 27.2 | 22.72 | NEG | Sx | | | | Y | | Y | | | | | | | 0 |
| 12 | F | POS | 35.74 | 25.98 | NEG | Sx | | Y | | | | | Y | | | | | | 1 |
| 14 | F | POS | 31.43 | 27.78 | NEG | Sx | Y | Y | Y | Y | Y | Y | Y | Y | | | | | 4 |
| 15 | F | POS | 34.2 | 27.7 | NEG | Sx | Y | | | | | | Y | | | | | | 1 |
| 1 | M | POS | 13.91 | 19.02 | POS | Sx | Y | | | | | | Y | | | | | | 6 |
| 1 | M | POS | 15.21 | 25.39 | POS | Sx | | | | | | Y | Y | Y | | | | | 1 |
| 7 | F | POS | 26.84 | 26.63 | POS | Sx | | Y | | | | | | | | | | | 2 |
| 9 | M | POS | 32.74 | 22.38 | POS | Sx | | | Y | Y | | Y | Y | | | | | Y | 1 |
| 9 | F | POS | 14.62 | 22.48 | POS | Sx | | Y | | Y | Y | | Y | | | | | | 2 |
| 10 | F | POS | 26.34 | 25.6 | POS | Sx | Y | | | | | | | | | | Y | | 2 |
| 10 | F | POS | 20.87 | 26.76 | POS | Sx | | | | Y | | | | Y | Y | | | | 1 |
| 11 | M | POS | 16.5 | 25.98 | POS | Sx | Y | Y | Y | | | | Y | Y | | | Y | Y | 2 |
| 11 | M | POS | 27.67 | 20.53 | POS | Sx | Y | | | | | | | Y | | | | | 4 |
| 12 | M | POS | 26.22 | 26.25 | POS | Sx | Y | Y | Y | Y | | Y | Y | Y | Y | | | | 5 |
| 14 | F | POS | 12.21 | 23.05 | POS | Sx | Y | Y | Y | Y | Y | | | Y | Y | | | | 3 |
| 14 | F | POS | 21.13 | 22.5 | POS | Sx | Y | Y | Y | Y | | | | | | | | | 2 |
| 14 | F | POS | 20.34 | 22.95 | POS | Sx | Y | | Y | Y | | Y | | Y | | | | | 2 |
| 14 | M | POS | 17.74 | 23.27 | POS | Sx | Y | Y | Y | | | Y | | Y | Y | | | | 7 |
| 16 | F | POS | 26.61 | 25.71 | POS | Sx | Y | Y | | Y | Y | | Y | | | | | Y | 6 |
| 17 | F | POS | 21.69 | 26.48 | POS | Sx | | Y | | | Y | | | Y | | | | Y | 4 |
| 17 | F | POS | 25.69 | 23.37 | POS | Sx | Y | Y | Y | | | Y | | | | | | Y | 0 |
| 18 | F | POS | 24.75 | 27.03 | POS | Sx | Y | | | | | | | | | | | | 1 |
| 18 | M | POS | 14.9 | 21.68 | POS | Sx | | Y | | Y | Y | | | | | | | | 1 |
| 18 | F | POS | 30.63 | 22.9 | POS | Sx | Y | | | Y | | | Y | Y | | | | | 1 |
| 18 | F | POS | 23.55 | 20.27 | POS | Sx | Y | | Y | Y | | Y | Y | Y | Y | | | Y | 4 |
| 18 | M | POS | 15.69 | 21.66 | POS | Sx | | Y | | Y | | | Y | Y | | | | | 1 |
| 2 | F | POS | 27.65 | 25.53 | POS | Sx7+ | | Y | | Y | | | | Y | | | Y | | 15 |
| 18 | F | POS | 26.79 | 27.71 | POS | Sx7+ | | Y | | Y | | | Y | Y | | | | Y | 11 |

F, female; M, male; POS, positive; Ct, cycle threshold; N2, SARS-CoV-2 target; RP, host control target; ASx, asymptomatic; Sx, symptomatic; n/a, not applicable

### Supplementary Table 3

### Distribution of positive and negative BinaxNOW and RT-PCR results across clinical subgroups

| Asymptomatic (Children: ≤18 yo) | | | |
|---|---|---|---|
| | RT-PCR | | |
| **BinaxNOW Ag** | Positive | Negative | Total |
| Positive | 70 | 7 | 77 |
| Negative | 37 | 715 | 752 |
| Total (12.9% prevalence) | 107 | 722 | 829 |

| Asymptomatic (Adults: >18 yo) | | | |
|---|---|---|---|
| | RT-PCR | | |
| **BinaxNOW Ag** | Positive | Negative | Total |
| Positive | 40 | 4 | 44 |
| Negative | 17 | 913 | 930 |
| Total (5.9% prevalence) | 57 | 917 | 974 |

| Symptomatic ≤7 days (Children: ≤18 yo) | | | |
|---|---|---|---|
| | RT-PCR | | |
| **BinaxNOW Ag** | Positive | Negative | Total |
| Positive | 22 | 0 | 22 |
| Negative | 4 | 65 | 69 |
| Total (28.6% prevalence) | 26 | 65 | 91 |

| Symptomatic ≤7 days (Adults: >18 yo) | | | |
|---|---|---|---|
| | RT-PCR | | |
| **BinaxNOW Ag** | Positive | Negative | Total |
| Positive | 82 | 0 | 82 |
| Negative | 3 | 270 | 273 |
| Total (23.9% prevalence) | 85 | 270 | 355 |

| Symptomatic (Children: >7 days) | | | |
|---|---|---|---|
| | RT-PCR | | |
| **BinaxNOW Ag** | Positive | Negative | Total |
| Positive | 2 | 0 | 2 |
| Negative | 0 | 6 | 6 |
| Total (25% Prevalence) | 2 | 6 | 8 |

| Symptomatic (Adults: >7 days) | | | |
|---|---|---|---|
| | RT-PCR | | |
| **BinaxNOW Ag** | Positive | Negative | Total |
| Positive | 10 | 1 | 11 |
| Negative | 5 | 35 | 40 |
| Total (29.4% prevalence) | 15 | 36 | 51 |

**Supplementary Table 4**

**Predictive value of the Abbott BinaxNOW for detection of SARS-CoV-2 in samples from adult and pediatric (≤18) patients, with varying prevalence, based on observed sensitivity/specificity in each subgroup.**

| | **Prevalence** | | | |
|---|---|---|---|---|
| | **20%** | **10%** | **5%** | **1%** |
| **PPV** | | | | |
| Adult, Sx ≤7D | 100% | 100% | 100% | 100% |
| Adult, ASx | 97.65% (93.91-99.12) | 94.87% (87.27-98.03) | 89.75% (76.45-95.93) | 62.68% (38.39-81.91) |
| Pediatric, Sx ≤7D | 100% | 100% | 100% | 100% |
| Pediatric, ASx | 93.41% (88.32-96.38) | 86.30% (77.06-92.20) | 74.91% (61.41-84.85) | 36.42% (23.40-51.80) |
| **NPV** | | | | |
| Adult, Sx ≤7D | 99.17% (98.38-99.58) | 99.63% (99.27-99.81) | 99.82% (99.65-99.91) | 99.97% (99.93-99.98) |
| Adult, ASx | 93.15% (90.11-95.30) | 96.83% (95.35-97.85) | 98.47% (97.74-98.97) | 99.70% (99.56-99.80) |
| Pediatric, Sx ≤7D | 96.30% (95.20-97.15) | 98.32% (97.81-98.71) | 99.20% (98.95-99.39) | 99.84% (99.80-99.88) |
| Pediatric, ASx | 91.89% (89.95-93.49) | 96.23% (95.27-97.00) | 98.18% (97.70-98.55) | 99.64% (99.55-99.72) |

PPV and NPV were calculated using the sensitivity and specificity observed in each subgroup for a hypothetical population of 1000 individuals per subgroup, with varying prevalence. PPV, positive predictive value; NPV, negative predictive value; Sx, symptomatic; ASx, asymptomatic; D, days

### Supplementary Table 5

#### Performance of the Abbott BinaxNOW versus RT-PCR with varied Ct value cutoffs

| | All Ct Values | | | | Ct values ≤ 35 | | | | Ct values ≤ 30 | | | | Ct values ≤ 25 | | | |
|---|---|---|---|---|---|---|---|---|---|---|---|---|---|---|---|---|
| | N | TP (+/+) | FN (+/-) | Sens | N | TP (+/+) | FN (+/-) | Sens | N | TP (+/+) | FN (+/-) | Sens | N | TP (+/+) | FN (+/-) | Sens |
| Combined All | 292 | 226 | 66 | 77.4% | 276 | 224 | 52 | 81.2% | 214 | 205 | 9 | 95.8% | 150 | 149 | 1 | 99.3% |
| All Children (Sx, ASx) | 135 | 94 | 41 | 69.6% | 125 | 92 | 33 | 73.6% | 92 | 83 | 9 | 90.2% | 62 | 61 | 1 | 98.4% |
| All Adult (Sx, ASx) | 157 | 132 | 25 | 84.1% | 151 | 132 | 19 | 87.4% | 122 | 122 | 0 | 100.0% | 88 | 88 | 0 | 100% |
| Sx (≤7D) Children | 26 | 22 | 4 | 84.6% | 25 | 22 | 3 | 88.0% | 21 | 20 | 1 | 95.2% | 14 | 14 | 0 | 100% |
| Sx (≤7D) Adult | 85 | 82 | 3 | 96.5% | 85 | 82 | 3 | 96.5% | 76 | 76 | 0 | 100.0% | 59 | 59 | 0 | 100% |
| ASx Children | 107 | 70 | 37 | 65.4% | 98 | 68 | 30 | 69.4% | 69 | 61 | 8 | 88.4% | 48 | 47 | 1 | 97.9% |
| ASx Adult | 57 | 40 | 17 | 70.2% | 51 | 40 | 11 | 78.4% | 37 | 37 | 0 | 100.0% | 24 | 24 | 0 | 100% |

Ct, cycle threshold; TP, true positive (RT-PCR positive/BinaxNOW positive); FN, false negative (RT-PCR positive/BinaxNOW negative); Sens, sensitivity; Sx, symptomatic; ASx, asymptomatic; D, days; Ped, pediatric

**Supplementary Table 6**

**Impact of temperature on BinaxNOW performance in individuals ≤7 days post symptom onset**

| **BinaxNOW result** | **RT-PCR result** | | | | | |
|---|---|---|---|---|---|---|
| | Temperature range: ≥59°F | | | Temperature range: 46-58.5°F | | |
| | Positive | Negative | Total | Positive | Negative | Total |
| Positive | 104 | 0 | 104 | 6 | 1 | 7 |
| Negative | 7 | 335 | 342 | 3 | 20 | 23 |
| Total | 111 | 335 | 446 | 9 | 21 | 30 |
| Sensitivity | (104/111) 93.7% | | | (6/9) 66.7% | | |
| Specificity | (335/335) 100.0% | | | (20/21) 95.2% | | |

The manufacturer recommends that tests be run at temperatures ≥ 59°F. Ag, antigen; Sx, symptoms

**Supplementary Figure 1**

**Correlation of Cycle Threshold with BinaxNOW band strength**

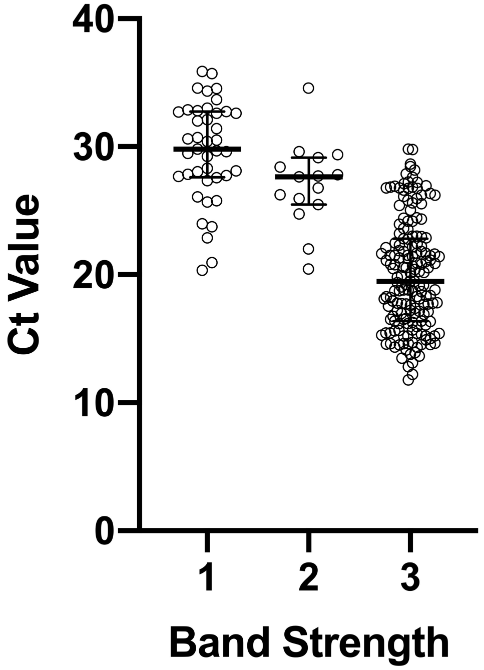


Band strength (1= faint, n = 41; 2 = medium, n = 15; 3 = strong, n = 170) as interpreted by the first reader for the 226 true positive BinaxNOW tests correlated clearly with Ct value, with median Cts of 29.7 (27.6-32.7), 27.7 (25.5-29.1), and 19.5 (16.4-22.8), respectively.
